# Prognostic significance of somatic mutations in myeloid cells of men with chronic heart failure – interaction between loss of Y chromosome and clonal hematopoiesis

**DOI:** 10.1101/2024.07.30.24310319

**Authors:** Sebastian Cremer, Moritz von Scheidt, Klara Kirschbaum, Lukas Tombor, Silvia Mas-Peiro, Wesley Abplanalp, Tina Rasper, Johannes Krefting, Akshay Ware, David Leistner, Heribert Schunkert, Thimoteus Speer, Stefanie Dimmeler, Andreas Michael Zeiher

**Affiliations:** Department of Medicine, Cardiology, Goethe University Hospital, Frankfurt, Germany; Institute for Cardiovascular Regeneration, Goethe University, Frankfurt, Germany; German Center for Cardiovascular Research, Berlin (partner site Frankfurt Rhine-Main), Germany; Cardiopulmonary Institute, Goethe University Frankfurt, Frankfurt, Germany; German Heart Center, Munich; Department of Medicine, Nephrology, Goethe University Hospital, Frankfurt, Germany; Else Kroener-Fresenius Center for Nephrological Research, Goethe University, Frankfurt, Germany

## Abstract

Age-associated clonal hematopoiesis of indeterminate potential (CHIP) has been associated with increased incidence and worse prognosis of chronic heart failure. CHIP is driven by somatic mutations in hematopoietic stem and progenitor cells (HSPC). Mosaic loss of the Y chromosome (LOY), the most common somatic mutation in blood cells of men, also correlates with clonal expansion of myeloid cells, increases with age and was experimentally shown to lead to diffuse cardiac fibrosis and subsequent heart failure in mice. However, the prognostic significance of LOY as well as its potential interaction with CHIP in patients with chronic heart failure is unknown. We investigated the prevalence and prognostic significance of the extent of LOY and the two most common CHIP-driver mutations DNMT3A and TET2 in 705 male patients with established chronic heart failure across the entire spectrum of left ventricular ejection fraction. Both, LOY and DNMT3A/TET2 mutations, increased with age, and LOY co-occurred with DNMT3A/TET2 mutations in 27.1% of men at age > 70 years. LOY was an independent predictor of death during 3-years of follow-up across the entire spectrum of left ventricular ejection fraction. The co-occurrence of harboring LOY and DNMT3A/TET2 mutations significantly contributed to the observed increased mortality observed in carriers of DNMT3A/TET2 mutations. The detrimental effect of LOY on prognosis was confirmed in a validation cohort of patients with ischemic heart disease. scRNA sequencing of peripheral blood cells in patients with chronic ischemic heart failure showed increased profibrotic signaling in LOY monocytes with elevated markers of monocyte mediated inflammation and profibrotic cardiac remodeling (S100A8, TLR2, CLEC4D) and reduced expression of TGF-β inhibiting genes (SMAD7, TGIF2). The proinflammatory phenotype of LOY monocytes was further amplified in LOY monocytes of patients simultaneously harboring DNMT3A mutations, who displayed heightened expression of alarmins (S100A8, HMGB2) and interferon signaling related genes (IFNGR1, TRIM56, CD84) compared to patients without CHIP mutations. Thus, the age-associated acquisition of somatic mutations in blood cells of men with chronic heart failure is associated with increased mortality, with loss of Y chromosome emerging as an independent predictor of all-cause death across the entire spectrum of left ventricular function.

## Introduction

Somatically acquired DNA mutations in hematopoietic stem cells have recently been identified as independent risk factors for the development and progression of cardiovascular disease.^1^ Most notably, clonal hematopoiesis of indeterminate potential (CHIP), defined as the presence of mutations in genes commonly associated with myeloid neoplasms in peripheral blood cells of subjects without any evidence for hematological malignancies, was previously shown to associate with the incidence and progression of chronic heart failure. ^2–4,5,6,7^. Specifically TET2 mutations also associate with incident heart failure with preserved ejection fraction, as just recently reported^8,9^. CHIP increases with age, confers a proliferative advantage leading to clonal expansion of mutant blood cells and is detected in >20% of patients with cardiovascular disease above the age of 70 years.^10,11,2^ On the other hand, mosaic loss of Y chromosome (LOY), defined as loss of the entire Y chromosome in a mosaic fashion, is the most common acquired somatic mutation in human blood cells and detected in >40% of men above the age of 70 years in the UK biobank.^12,13^ Similar to CHIP, LOY increases with age and correlates with clonal expansion of myeloid cells.^12,14^ Most importantly, Sano et al.^15^ recently experimentally demonstrated that hematopoietic loss of Y chromosome in mice led to diffuse cardiac fibrosis during aging culminating in the development of heart failure.

We could recently demonstrate that LOY in blood cells is associated with profoundly impaired long-term survival even after successful removal of the stenotic aortic valve in men undergoing TAVR.^16^ However, the prognostic significance of LOY in established chronic heart failure has not been assessed so far. Moreover, given the similarities in age-related prevalence and cardiovascular disease risk, it is tempting to speculate that LOY and CHIP may represent two sides of the same coin.^17^

Therefore, in the present study, we investigated both, the prognostic significance of LOY as well as its potential interaction with CHIP in men with chronic heart failure.

## Methods

### Study cohort

In this institutional review board approved study, clinical data and peripheral blood cells were collected from a total of 705 male patients with established heart failure. Patients were eligible for inclusion into the present analysis, if they suffered from stable chronic heart failure symptoms NYHA ≥2, clinical evidence of heart failure and elevated NT-pro BNP serum levels. Exclusion criteria were the presence of acutely decompensated heart failure with NYHA class IV, an ischemic cardiac event or interventional procedure within 30 days prior to inclusion, documented hematological disease or cancer within the preceding 5=years, or unwillingness to participate. The CHIP results of some of the patients with chronic ischemic heart failure and reduced LVEF have been previously reported.^2^ Coronary artery disease was defined as prior myocardial infarction or revascularization. In addition, some of the patients reported in our most recent study describing a prognostic significance of LoY after TAVR^16^ were also included, if they suffered from continuous heart failure symptoms NYHA ≥ 2 and elevated NTpro-BNP serum levels at least 30 days after successful valve replacement. The primary endpoint of the study was all-cause mortality during a prospective 3 years of follow-up.

Replication of results was performed in an independent external validation cohort at the German Heart Center Munich. Following the same criteria, 2,003 patients with confirmed coronary artery disease (CAD), who underwent coronary angiography between January 2014 and December 2022, were screened for LoY and CHIP status. Propensity score matching (1:1) was employed to minimize potential bias between comparison groups based on most relevant clinical variables and risk factors.

The study complied with the requirements of the Declaration of Helsinki. All participants provided written informed consent for the study procedures, including genetic testing using blood samples.

### Measurement of LOY

Estimation of LOY was performed using a previously validated digital PCR technique.^14^ In brief, the TaqMan-based method quantifies the relative number of X and Y chromosomes in a DNA sample by targeting a 6 bp sequence difference present between the AMELX and AMELY genes using the same primer pair and, thus, is relatively unbiased with regard to primer properties. DNA was extracted from whole blood at the timepoint of study inclusion.

As previously described,^16^ 150 nanograms of DNA was mixed with Probe PCR Master Mix (QIAcuity Probe PCR Kit, #250101, Qiagen, Hilden, Germany) containing FastDigest HindIII enzyme (#FD0504, ThermoFisher Scientific, Waltham, MA, USA) and TaqMan Primers (#C_990000001_10, ThermoFisher Scientific, Waltham, MA, USA). After digestion for 10 min at room temperature, a 26k 24-well Nanoplate (QIAcuity Nanoplate 26k 24-well, #250001, Qiagen, Hilden, Germany) was filled with the reaction mix and loaded into a QIAcuity One instrument (Qiagen, Hilden, Germany). PCR cycling was performed following manufacturer’s instructions: PCR initial heat activation 95°C for 2 min, followed by two-step cycling (40 cycles) of 95°C for 15 s and 60°C for 30 s. A 6 nt deletion occurs in the X-specific *amelogenin* gene (B37/hg19 genome locations: chrX:11315039 and chrY:6 737 949– 6 737 954). The VIC dye probe detects X-chromosome sequences, and the FAM dye probe includes the 6nt and detects Y-chromosome sequences (Sequence: GTGTTGATTCTTTATCCCAGATG[-/<u>AAGTGG</u>]TTTCTCAAGTGGTCCTGATTTT [VIC/FAM]).

The endpoint fluorescence intensity of the partitions was separately measured for FAM (targeting AMELY) and VIC (targeting AMELX) to determine the presence or absence of the respective targets. Using the QIAcuity One Software Suite (Qiagen, Hilden, Germany) the absolute concentration of the targets was calculated based on the number of positive and negative partitions. The ratio of AMELY/AMELX was calculated using absolute concentrations. The extent of LOY as percentage was defined as the ratio of AMELY copy numbers in relation to AMELX copy numbers and calculated by the formula: 1-Copy numbers AMELY / Copy Numbers AMELX)*100

### Sequencing for DNMT3A and TET2-mediated clonal hematopoiesis

The presence of somatic *DNMT3A* or *TET2* CHIP-driver mutations with a variant allele frequency (VAF) ≥ 2% in patients with aortic stenosis as well as in the replication cohort was commercially assessed by next generation sequencing (NGS), as extensively described previously. ^18,19^ In brief, the patients’ libraries were generated with the Nextera Flex for enrichment kit (Illumina, San Diego, CA, USA) and sequences for DNMT3A and TET2 enriched with the IDT xGen hybridisation capture of DNA libraries protocol and customised probes (IDT, Coralville, IA). The libraries were sequenced on an Illumina NovaSeq 6000 with a mean coverage of 2147 × and a minimum coverage of 400×, reaching a sensitivity of 2%. Reads were mapped to the reference genome (UCSC hg19) using Isaac aligner (v2.10.12) and a small somatic variant calling was performed with Pisces (v5.1.3.60). Protein truncating variants were classified as mutation. Non-synonymous changes were included, if they were well annotated (several definite submissions to COSMIC, IRAC or ClinVAR). Other non-protein truncating variants were defined as variants of uncertain significance (VUS).

In the patients with ischemic HFrEF, a custom panel based on the Illumina TruSeq Custom Amplicon Low Input assay was designed to assess the presence of CHIP, including 594 amplicons in 56 genes commonly mutated in CHIP and myeloid malignancies. The pooled libraries were sequenced on a NextSeq 500 sequencer (Illumina, USA) using the NextSeq 500/550 Mid Output v2 kit (300 cycles) according to the manufacturer’s instructions. The median coverage across all samples was 4282x before UMI family clustering and 630x with inclusion of UMIs resulting in a sensitivity of 0.5% ^2^.

CHIP variants for DNMT3A and TET2 can be found in tables **Supplementary Table 1** and **Supplementary Table 2**, respectively.

### Single cell RNA sequencing (sc-RNAseq) analyses

Sc-RNAseq was performed as previously described ^20^. In brief, patient-derived blood was centrifuged on density gradient centrifugation (Pancoll human, Density: 1.077 g/mL, #P04-60500, PAN-Biotech GmbH, Aidenbach, Germany), and mononuclear cells were used for droplet scRNAseq using the Chromium Controller with Chromium Next GEM Single Cell 3′ GEM, Library and Gel Bead Kit version 3.1 reagent (10 × Genomics, Pleasanton, CA) according to the manufacturer’s protocol. Libraries were sequenced using paired-end sequencing by GenomeScan (Leiden, Netherlands), and expression data were processed by the Cell Ranger Single Cell Software Suite version 3 (10 × Genomics, Pleasanton, CA, USA) and aligned to the human reference genome GRCh38. Data integration was performed by Seurat version (v4.0.3) (Satija Lab, New York Genome Center, New York City, NY, USA), and FindMarkers function in the Seurat package was used for statistical analysis of differential gene expression. Loss of Y chromosome cells were defined in scRNAseq data as combinatorial lack of expression of all Y chromosome derived genes. Relative changes in transcription of Y harboring and LOY cells were then assessed and data were pooled. For subsequent analyses, relative changes in transcription of Y harboring and LOY cells within each patient were assessed, allowing for paired analyses in patients with DNMT3A driver mutations and without CHIP mutations.

### Statistical analyses

Categorical variables are presented as numbers and frequencies (%), and continuous variables as median [interquartile range (IQR)] unless otherwise noted. We used Cochran-Armitage tests for trend to compute significance values for linear trends of categorical variables over increasing LoY quartiles. Continuous variables were compared between groups by the Wilcoxon rank sum test. The best cut-off value for the extent of LOY in predicting mortality was calculated using the Youden index^21^ based on the area under the curve (AUC) from a smooth receiver operating characteristic (ROC) curve estimation for right censored data. Confidence intervals and standard errors were estimated with a bootstrapping approach ^22^. For mortality analysis, LOY data were dichotomized according to the cut-off value to define the groups of patients with high vs. low LOY. Three-year survival was displayed as Kaplan–Meier plots with log-rank statistic. Univariate and multivariate Cox regression analyses were used to model predictor influences on mortality. All covariables reaching a *P*-value <0.05 in univariate analysis were included in multivariate analysis to adjust for potential confounding effects. HR plots for the association between LOY and CHIP with mortality according to baseline EF were built using STATA IC 15.0. Cause-specific survival was modeled using flexible parametric models included in the STATA package ‘stpm2’. Restricted cubic splines of EF were built using the STATA package ‘rcsgen’ with three degrees of freedom (knots at 0^th^, 33^rd^, 67^th^, and 100^th^ centile). The final model included the splines of EF, presence of LOY or CHIP, as well as interaction terms between LOY/CHIP and the splines of EF as independent variables. Data analysis was performed with R, version 4.2.1.

Sc-RNAseq analysis was performed by using R (version 4.0.3) and the analysis tool Seurat (v4.0.3). In brief, differential expression of genes was used utilizing the ‘FindMarkers’ function in the Seurat package for focused analyses. Significant comparisons were performed using the Wilcoxon Rank Sum test followed by Bonferroni correction. Significance was reported if adjusted *P* < 0.05. Differential transcriptional profiles by LOY status were generated in Seurat with associated gene ontology terms derived from the functional annotation tool Metascape (v3.5) for further analyses.

## Results

### Patient characteristics

The clinical characteristics and laboratory values of the patient cohort are summarized in **Table 1**. Patients had a mean age of 75 (62-82) years. The extent of LOY in blood cells ranged from -13.3% to 83.4%. 207 patients (29.4%) had LOY ≥ 10%. Calculation of the Youden-Index revealed an optimal cut-off value for LOY of 17% to predict mortality during three years of follow-up. 131 patients (18.6%) had LOY ≥ 17%. A total of 82 patients (12%) had DNMT3A CHIP-driver mutations with a VAF ≥ 2%, 71 patients (11%) had TET2 mutations with a VAF ≥ 2%, and 9 patients had both DNMT3A and TET2 CHIP-driver mutations. Altogether, 144 patients harbored DNMT3A and/or TET2 CHIP-driver mutations. The median VAF of patients with DNMT3A mutations was 5.33% (IQR 3.4%;12.7%) the median VAF of patients with TET2 mutations was 5.4% (IQR 3.3%;11%).

**Table 1.** Baseline characteristics of all patients and patients with LOY < 17% compared to ≥17%. Continuous variables are presented as median with interquartile range.

Baseline
| Characteristic | N = 705 | LOY < 17%<br>N = 574 | LOY≥17%<br>N=131 | p-value |
| --- | --- | --- | --- | --- |
| Cohort |  |  |  |  |
| Ischemic Heart Failure | 333 (47%) | 308 (54%) | 25 (19%) | <0.001 |
| TAVR | 372 (53%) | 266 (46%) | 106 (81%) |  |
| DNMT3A VAF≥2 | 82 (12%) | 66 (11%) | 16 (12%) | 0.8 |
| TET2 VAF≥2 | 71 (10%) | 50 (8.7%) | 21 (16%) | 0.012 |
| DNMT3A and or TET2 VAF ≥2 | 144 (20.3%) | 109 (19.2%) | 35 (27%) | 0.048 |
| COPD (N=701) | 81 (12%) | 64 (11%) | 17 (13%) | 0.6 |
| Diabetes mellitus (N=702) | 216 (31%) | 181 (32%) | 35 (27%) | 0.3 |
| Coronary Artery Disease (N=701) | 585 (83%) | 485 (85%) | 100 (76%) | 0.015 |
| Hypertension (N=702) | 543 (77%) | 442 (77%) | 101 (77%) | >0.9 |
| Atrial Fibrillation (N=695) | 403 (58%) | 331 (59%) | 72 (55%) | 0.4 |
| History of Smoking (N=468) | 309 (66%) | 264 (66%) | 45 (66%) | >0.9 |
| Age | 75 (62 ,82) | 72 (59, 80) | 81 (75, 83) | <0.001 |
| BMI (N=697) | 26.8 (24.6, 29.4) | 27.0 (24.6, 29.4) | 26.0 (24.3, 28.9) | 0.072 |
| LVEF (%) | 40 (30, 55) | 40 (29, 55) | 50 (35, 60) | <0.001 |
| LVEF <50% | 436 (62%) | 373 (65%) | 63 (48%) |  |
| LVEF ≥ 50% | 269 (38%) | 201 (35%) | 68 (52%) |  |
| Creatinine (N=699) | 1.13 (0.96, 1.41) | 1.12 (0.95, 1.36) | 1.20 (0.96, 1.57) | 0.12 |
| CRP (N=647) | 0.30 (0.14, 0.86) | 0.31 (0.15, 0.84) | 0.26 (0.10, 0.92) | 0.3 |
| Hemoglobine (g/dl); N=701 | 13.60 (12.00, 14.80) | 13.80 (12.12, 14.97) | 12.60 (11.00, 14.00) | <0.001 |
| Thrombocytes (/nl); N=650 | 202 (166, 247) | 201 (167, 247) | 206 (166, 244) | 0.8 |
| Leukocytes (/nl); N=700 | 7.24 (6.04, 8.78) | 7.30 (6.08, 8.81) | 6.96 (5.95, 8.28) | 0.065 |
| hs Troponin (pg/ml); N=372 | 26 (15, 47) | 25 (15, 49) | 27 (16, 42) | >0.9 |
| NT-proBNP (pg/ml); N=486 | 1441 (539, 3266) | 1311 (488, 3007) | 1702 (792, 3954) | 0.02 |

### Prevalence of LOY and DNMT3A/TET2 CHIP driver mutations

As illustrated in **Figure 1A**, both LOY and DNMT3A/TET2 CHIP driver mutations increased with age. Above the age of 70 years, 116 patients (27.2%) showed LOY ≥17%, whereas 122 patients (28.6%) carried DNMT3A and/or TET2 CHIP-driver mutations. Importantly, the prevalence of harboring DNMT3A/TET2 CHIP-driver mutations significantly increased from 12.4% in the patients in the lowest quartile of the extent of LOY up to 25% in the highest quartile of the extent of LOY **(Figure 1B)**. When characterizing the 131 patients with LOY ≥ 17%, 35 patients (27%) simultaneously harbored DNMT3A/TET2 CHIP-driver mutations. There was no significant correlation between the extent of LOY and the VAF for both, DNMT3A and TET2 CHIP-driver mutations **(Supplementary** Figure 1).

**Figure 1.**
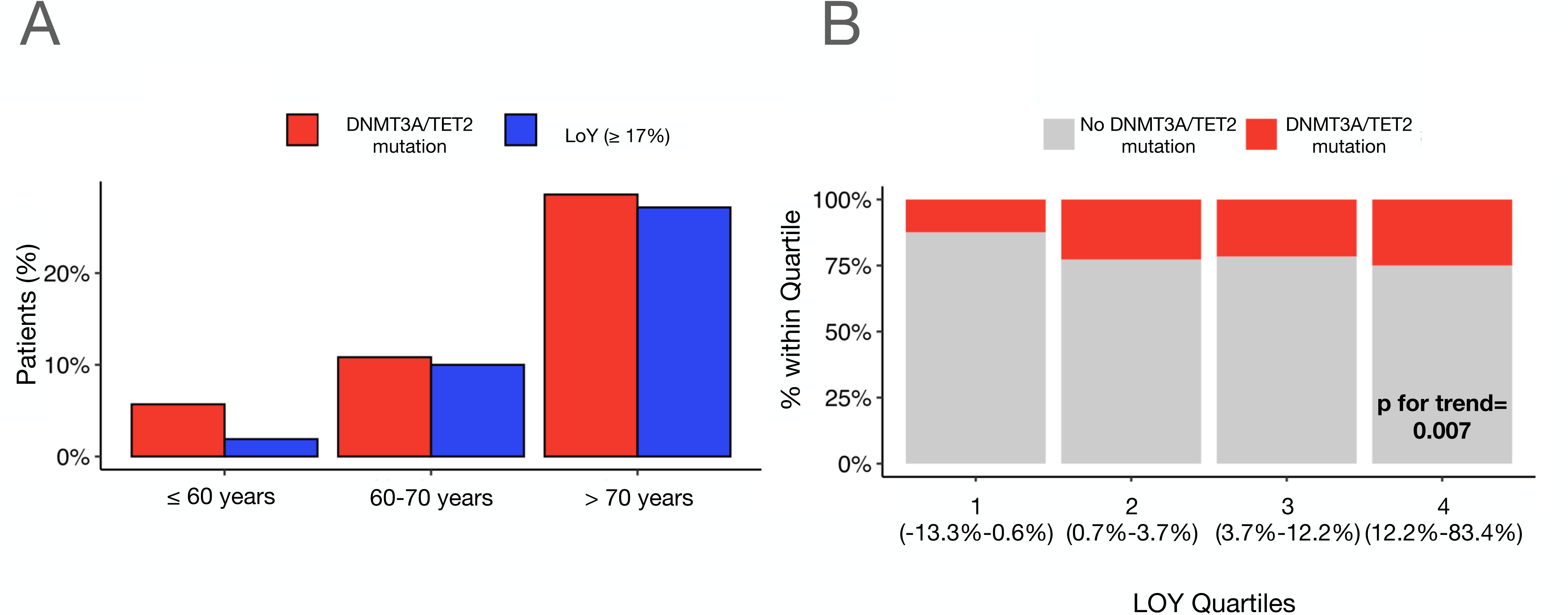
**(A)** Prevalence of LOY ≥17% and DNMT3A/TET2 CHIP-driver mutations according to age. **(B)** Prevalence of DNMT3A/TET2 CHIP-driver mutations in LOY quartiles. The range of LOY within each quartile is indicated. Cochran-Armitage test for trend (Z=3.12, p=0.002).

### Prognostic significance of LOY and DNMT3A/TET2 CHIP driver mutations

The clinical characteristics and laboratory parameters of the 131 patients (18.6% of the whole cohort) with LOY ≥ 17% are compared with those of the patients with LOY < 17% in **Table 1**. As expected, patients with LOY ≥ 17% were significantly older, but also had significantly higher NT-pro BNP serum levels and higher values of left ventricular ejection fraction (LVEF) as well as lower hemoglobin levels and a slightly reduced incidence of coronary artery disease (CAD), while the prevalence of DNMT3A/TET2 driver mutations was significantly increased. All other measured parameters including kidney function, COPD and diabetes did not differ. There was a dose dependent significant increase in mortality with increasing extent of LOY ranging from 11.9% in the lowest quartile to 23.3% in the highest quartile of LOY **(Figure 2A).** Moreover, as illustrated in **Figure 2B**, Cox regression analysis revealed that LOY as a continuous variable is a significant predictor of death in patients with heart failure (HR 1.02, CI 1.01-1.03; p=0.004) with a sharp increase in HR for death at LOY≥ 17% **(Figure 2B).**

**Figure 2.**
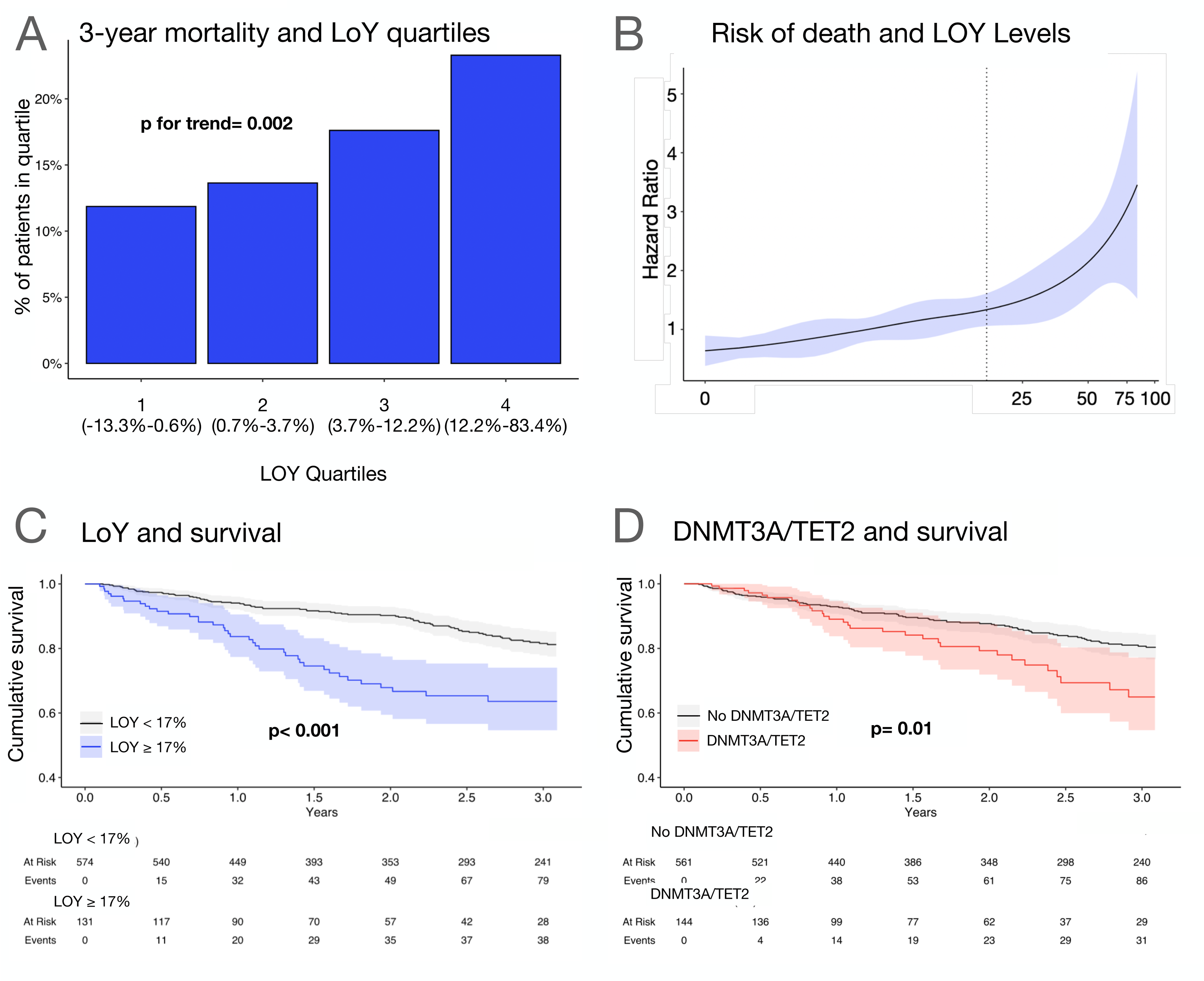
**(A)** Three-year mortality according to LOY quartiles. Increase in mortality was significant. The range of LOY within each quartile is indicated. Cochran-Armitage test for trend (Z=3.054, p=0.002). **(B)** Extent of LOY as continuous variable and hazard ratios for all-cause mortality according to a cubic spline regression model, shaded areas represent 95% confidence intervals, dashed line denotes LOY = 17%. **(C)** Kaplan-Meier survival curves in patients with LOY≥17% compared to patients with LOY < 17%. **(D)** Kaplan-Meier survival curves of patients harboring DNMT3A/TET2 CHIP-driver mutations compared to patients without DNMT3A/TET2 CHIP-driver mutations.

During three years of follow up, 38 of 131 patients (29%) showing LOY ≥ 17% compared to 79 of 574 patients (13.8%) with LOY < 17% died. **Figure 2C** illustrates the Kaplan-Meier survival curves of the 2 cohorts demonstrating the significantly increased mortality in men having ≥ 17% LOY in circulating blood cells. Similarly, as previously reported^2,4^, harboring a DNMT3A/TET2 CHIP driver mutation with a VAF ≥ 2% conferred an increased risk of death during follow up **(Figure 2D).**

However, on multivariate analysis including all differentially distributed baseline characteristics (NT-pro BNP, age, LVEF, hemoglobin, and coronary artery disease), in addition to age, LVEF, NTproBNP and hemoglobin levels, LOY ≥ 17%, but not the presence of DNMT3A/TET2 CHIP driver mutations remained a significant independent predictor of mortality during follow up (**Figure 3A).** Thus, LOY ≥ 17% appears to be independently associated with increased mortality during follow-up irrespective of the presence of DNMT3A/TET2 CHIP driver mutations. In order to precisely assess the relative contribution of harboring LOY and DNMT3A/TET2 CHIP-driver mutations, we calculated the hazard ratios for mortality for the individual patient groups with the patients with LOY < 17% and no DNMT3A/TET2 CHIP-driver mutation set as comparator. As illustrated in **Figure 3B**, harboring only DNMT3A/TET2 CHIP-driver mutations, but no LOY ≥ 17% was associated with a HR of 1.52 (95% CI 0.93-2.49), while harboring LOY ≥ 17%, but no DNMT3A/TET2 CHIP-driver mutation resulted in a HR of 2.34 (95% CI 1.52-3.62) and the presence of both, LOY ≥ 17% and DNMT3A/TET2 CHIP-driver mutations, gave the highest HR for mortality with 3.67 (95% CI 2.03-6.62). Statistical analysis demonstrated a significant interaction (p<0.001) between the different HR indicating a significant effect of harboring LOY ≥ 17% over and above DNMT3A/TET2 CHIP-driver mutations on mortality. Multivariate analysis including nt-proBNP, LVEF, hemoglobin levels, thrombocyte numbers, presence of coronary artery disease, age, and hs-CRP confirmed that the presence of DNMT3A/TET2 mutations in conjunction with LOY ≥17% is associated with the highest risk for mortality in patients with heart failure (HR 2.75 (95% CI 1.32-5.74; p=0.007, **Figure 3C**).

**Figure 3.**
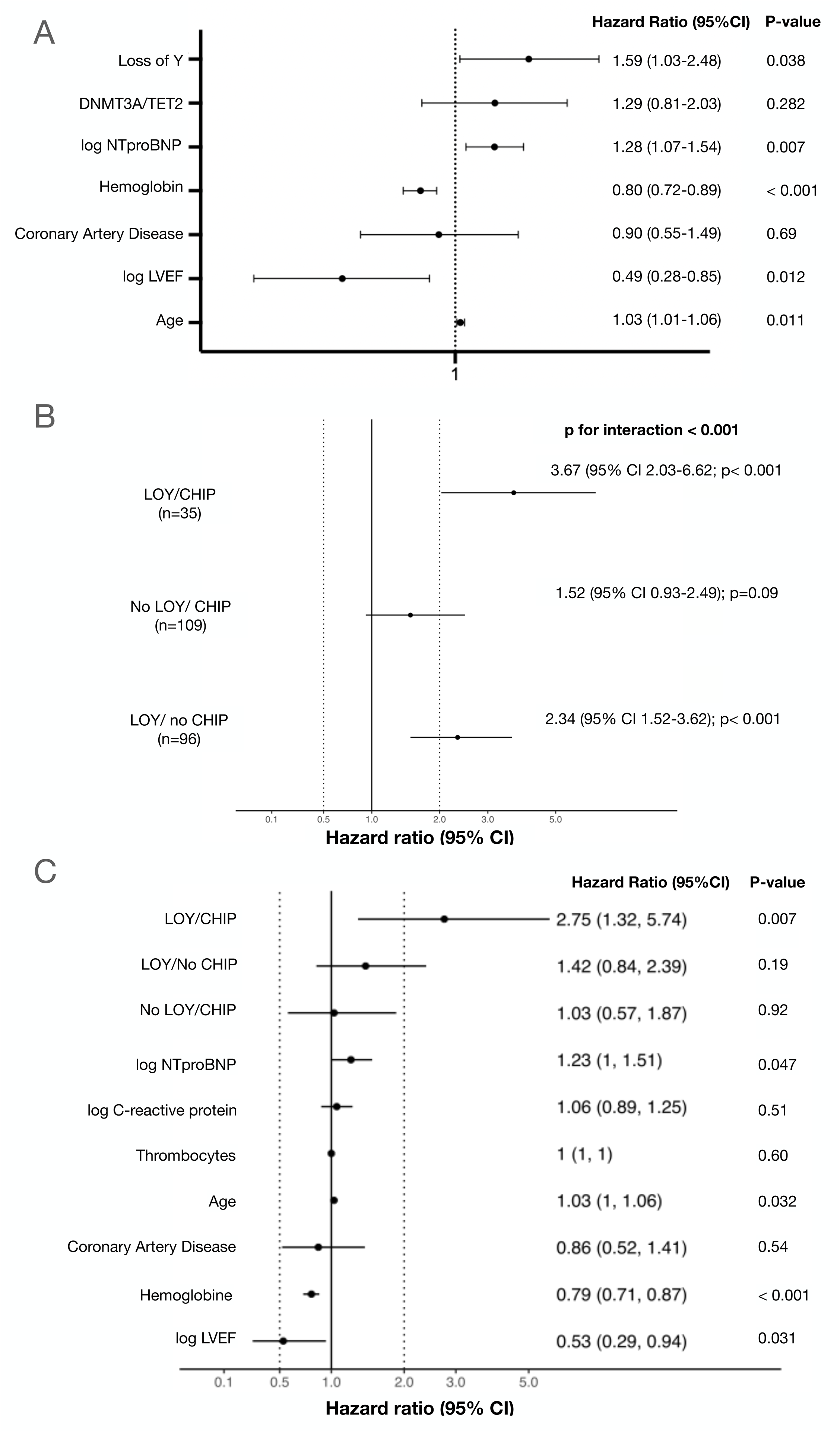
**(A)** Multivariate analysis of all-cause mortality. Hazard ratio (HR) and 95% confidence intervals (CI) are displayed. **(B)** Univariate hazard ratios for all-cause mortality for different groups according to the presence of either LOY or DNMT3/TET2 CHIP or both, LOY and DNMT3/TET2 CHIP. Hazard ratio (HR) and 95% confidence intervals (CI) are displayed. N-numbers for each group are indicated. 461 men neither harbored LOY ≥ 17% nor DNMT3A/TET2 CHIP-driver mutations. **(C)** Multivariate analysis of all-cause mortality. Hazard ratio (HR) and 95% confidence intervals (CI) are displayed.

Importantly, there were no significant differences in clinical characteristics or laboratory parameters in DNMT3A/TET2 CHIP driver mutation carriers with or without LOY ≥ 17% (**Table 3).**

**Table 2.** Baseline characteristics of all patients and patients with DNMT3A/TET2 mutations compared to patients without DNMT3A/TET2 mutations. Continuous variables are presented as median with interquartile range.

Baseline
| Characteristic | N = 705 | No DNMT3A/TET2<br>N = 561 | DNMT3A/TET2<br>N=144 | p-value |
| --- | --- | --- | --- | --- |
| Cohort |  |  |  |  |
| Ischemic Heart Failure | 333 (47%) | 303 (54%) | 30 (21%) | <0.001 |
| TAVR | 372 (53%) | 258 (46%) | 114 (79%) |  |
| DNMT3A VAF≥2 | 82 (12%) | 0 (0%) | 82 (57%) | <0.001 |
| TET2 VAF≥2 | 71 (10%) | 0 (0%) | 71 (49%) | <0.001 |
| DNMT3A and or TET2 VAF ≥2 | 144 (20.3%) | 0 (0%) | 144 (100%) | <0.001 |
| LoY |  |  |  |  |
| LoY <17% |  | 465 (83%) | 109 (76%) | 0.048 |
| LoY ≥ 17% |  | 96 (17%) | 35 (24%) |  |
| COPD (N=701) | 81 (12%) | 63 (11%) | 18 (13%) | 0.6 |
| Diabetes mellitus (N=702) | 216 (31%) | 165 (29%) | 51 (36%) | 0.14 |
| Coronary Artery Disease (N=701) | 585 (83%) | 472 (84%) | 113 (80%) | 0.2 |
| Hypertension (N=702) | 543 (77%) | 429 (77%) | 114 (80%) | 0.4 |
| Atrial Fibrillation (N=695) | 403 (58%) | 331 (60%) | 72 (51%) | 0.062 |
| History of Smoking (N=468) | 309 (66%) | 260 (67%) | 49 (62%) | 0.4 |
| Age | 75 (62 ,82) | 73 (60, 80) | 80 (74, 84) | <0.001 |
| BMI (N=697) | 26.8 (24.6, 29.4) | 26.8 (24.6, 29.4) | 26.6 (24.5, 29.1) | >0.9 |
| LVEF (%) | 40 (30, 55) | 40 (29, 55) | 50 (38, 60) | <0.001 |
| LVEF <50% | 436 (62%) | 367 (65%) | 69 (48%) |  |
| LVEF≥ 50% | 269 (38%) | 194 (35%) | 75 (52%) |  |
| Creatinine (N=699) | 1.13 (0.96, 1.41) | 1.13 (0.97, 1.41) | 1.13 (0.93, 1.39) | 0.7 |
| CRP (N=647) | 0.30 (0.14, 0.86) | 0.31 (0.16, 0.86) | 0.21 (0.09, 0.79) | 0.014 |
| Hemoglobine (g/dl); N=701 | 13.60 (12.00, 14.80) | 13.80 (12.00, 15.00) | 13.00 (11.20, 14.17) | <0.001 |
| Thrombocytes (/nl); N=650 | 202 (166, 247) | 206 (169, 249) | 186 (162, 232) | 0.031 |
| Leukocytes (/nl); N=700 | 7.24 (6.04, 8.78) | 7.28 (6.09, 8.79) | 6.98 (5.88, 8.66) | 0.2 |
| hs Troponin (pg/ml); N=372 | 26 (15, 47) | 26 (15, 48) | 26 (15, 47) | >0.9 |
| NT-proBNP (pg/ml); N=486 | 1441 (539, 3266) | 1,418 (518, 2,945) | 1,594 (616, 4,892) | 0.4 |

**Table 3.**
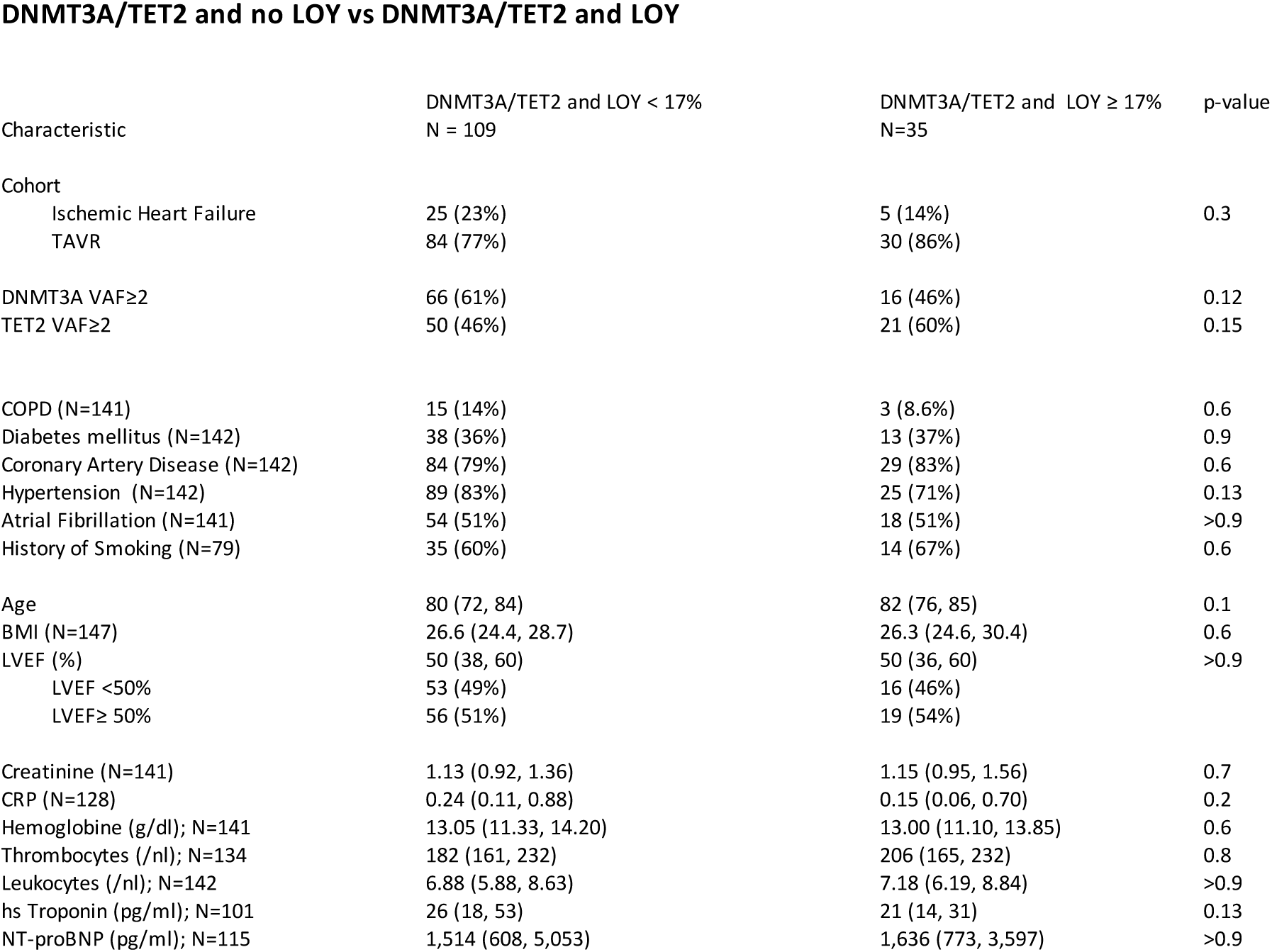
Clinical characteristics of DNMT3A/TET2 CHIP-driver mutation carriers with LOY <17% compared to DNMT3A/TET2 CHIP-driver mutation carriers with LOY ≥ 17%. Continuous variables are presented as median with interquartile range.

Thus, LOY ≥ 17%, which co-occurs with DNMT3A/TET2 CHIP driver mutations in more than 20% of all cases studied and in 27.2 % of all patients at the age above 70 years, appears to be a major determinant of increased mortality in DNMT3A/TET2 CHIP driver mutation carriers with chronic heart failure.

### Prognostic significance of LOY and DNMT3A/TET2 CHIP driver mutations across the entire spectrum of LVEF

To assess whether there is an interaction between harboring LOY ≥17% or DNMT3A/TET2-CHIP driver mutations with LVEF with respect to the mortality HR, we constructed cubic spline functions relating HR for mortality to LVEF across the entire spectrum of LVEF. As illustrated in **Figure 4A**, LOY ≥ 17% is an independent predictor of increased mortality (HR including confidence intervals > 1) across the entire spectrum of LVEF, while harboring DNMT3A/TET2 CHIP-driver mutations were only significantly associated with a statistically significant increase in the HR for mortality in patients with a baseline LVEF < 45% (**Figure 4B**). This is further illustrated by dichotomizing the patients according to a baseline LVEF < 50% or ≥ 50%, which is the classical definition of reduced versus preserved LVEF and a threshold for changing patient characteristics in heart failure^23,24^. For patients with a baseline LVEF < 50%, both, harboring LOY ≥ 17% as well as DNMT3A/TET2 CHIP-driver mutations, were significantly associated with increased all-cause mortality by Kaplan-Meier curve analysis (**Figure 4C/D**), while only the presence of LOY ≥ 17%, but not DNMT3A/TET2 CHIP-driver mutations was associated with a significantly worse long-term outcome in patients with a baseline LVEF ≥ 50% (**Figure 4E/F**). Similar results were obtained when hazards were calculated independently in subgroups of patients divided into the guideline-recommended groups of reduced LVEF ≤40% (HFrEF), mildly reduced LVEF 41-49% (HFmrEF), and preserved LVEF ≥ 50% (HFpEF). Here, harboring LOY ≥ 17% was associated with a statistically significant increase in mortality in all groups, while harboring DNMT3A/TET2 CHIP-driver mutations was only a significant factor in men with HFrEF, but not with HFmrEF or HFpEF (**Supplementary** Figure 2).

**Figure 4.**
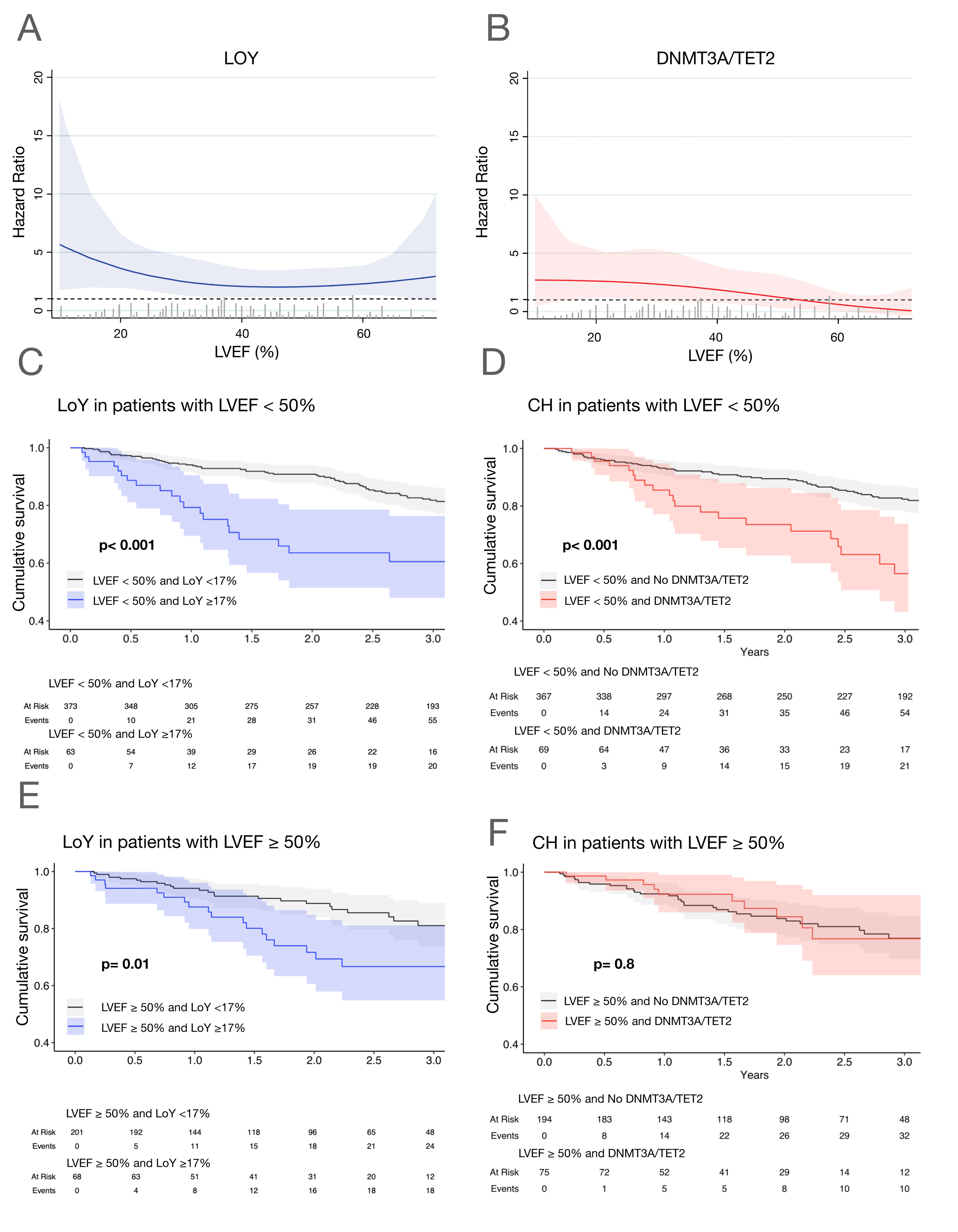
**(A)** Influence of LVEF on the effect of harboring LOY ≥ 17% on mortality. LVEF is analyzed as a continuous variable and the relationship is depicted according to a cubic spline regression model. Shaded areas represent 95% confidence intervals. **(B)** Influence of LVEF on the effect of harboring a DNMT3A/TET2 CHIP-driver mutation on mortality. LVEF is analyzed as a continuous variable and the relationship is depicted according to a cubic spline regression model. Shaded areas represent 95% confidence intervals. **C** Kaplan-Meier survival curves in patients with LOY≥17% compared to patients with LOY < 17% in patients with LVEF < 50% **D** Kaplan-Meier survival curves in patients harboring DNMT3A/TET2 CHIP-driver mutations versus patients without DNMT3A/TET2 CHIP-driver mutations with LVEF < 50%. **E** Kaplan-Meier survival curves in patients with LOY≥17% compared to patients with LOY < 17% in patients with LVEF ≥ 50% **F** Kaplan-Meier survival curves in patients harboring DNMT3A/TET2 CHIP-driver mutations versus patients without DNMT3A/TET2 CHIP-driver mutations with LVEF ≥ 50%.

Finally, we validated the main findings of our study in an independent validation cohort of a total of 2003 male patients undergoing coronary angiography at the Munich Heart Centre. In this slightly younger cohort, 324 patients (16,2 %) had LOY ≥ 17%, but no DNMT3A or TET2 mutations, and 261 of those patients had a LVEF <45% and/or two- or three-vessel coronary artery disease. In order to exclude potential confounding factors, the 261 patients with LOY ≥17%, but without DNMT3A/TET2 mutations were matched in a 1:1 fashion with 261 patients with LOY < 17% and absent DNMT3A/TET2 mutations. Matching was performed for age, affected coronary arteries, LVEF, arterial hypertension, smoking, and history of myocardial infarction **(Figure 5A).** As illustrated in **Figure 5B**, patients with LOY ≥17%, but no DNMT3A/TET2 mutations had a significantly increased mortality during a three-year follow-up with a hazard ratio of 2.10 (1.04 – 4.23; p=0.04). In contrast, for 179 patients with DNMT3A and/or TET2 mutations, but LOY < 17% matched in a similar fashion **(Figure 5C)**, the presence of DNMT3A/TET2 mutations did not confer a significantly increased risk of death in the absence of LOY < 17% **(Figure 5D).** Thus, the results of this validation cohort confirm that LOY is an independent predictor of death in patients with ischemic heart disease.

**Figure 5.**
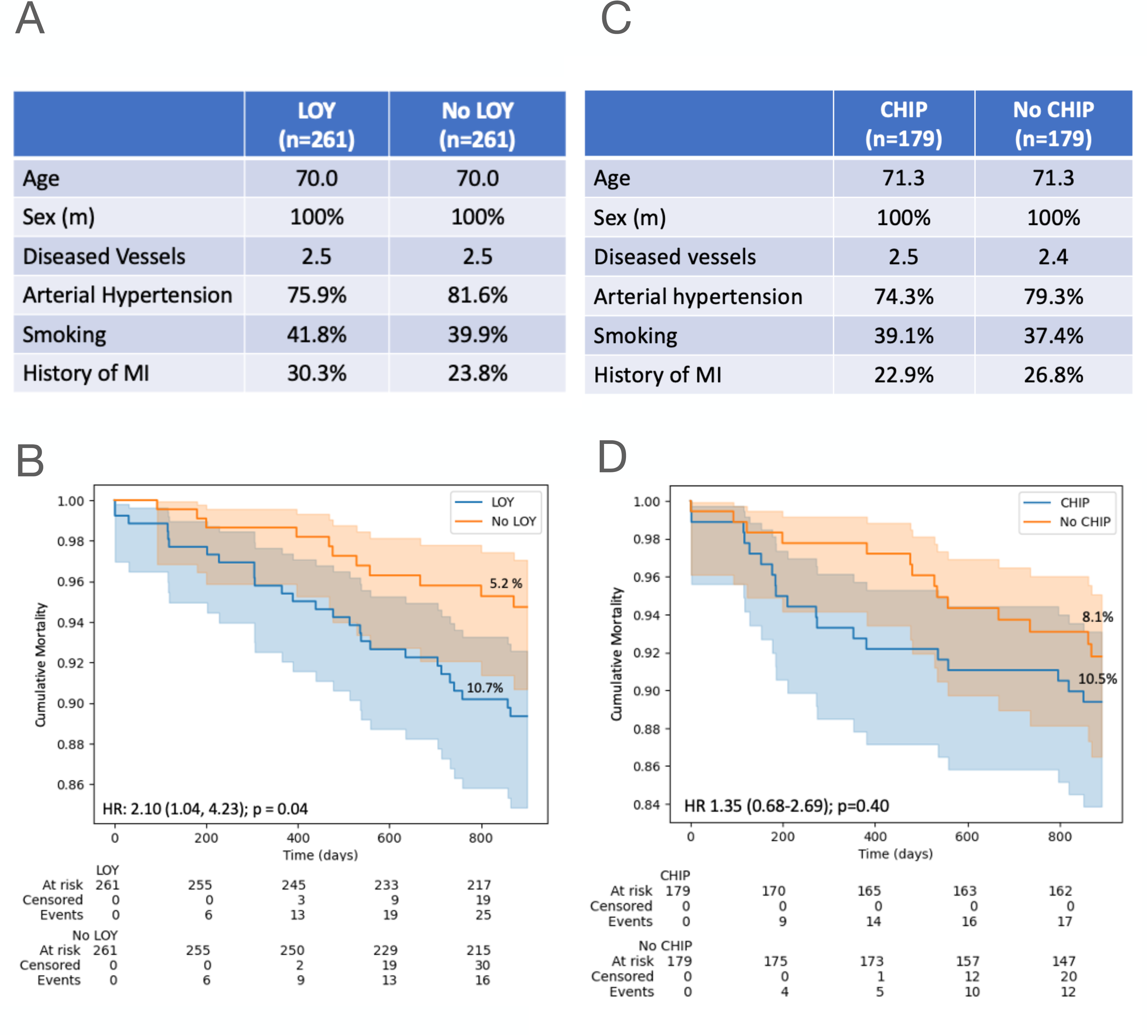
**(A)** Key matching parameters of the 261 patients with and without LOY ≥ 17%, but absent DNMT3A/TET2 CHIP-driver mutations. **(B)** Kaplan-Meier survival curves in patients with absent DNMT3A/TET2 CHIP-driver mutations, but with LOY ≥ 17%, compared to LOY <17%. **(C)** Key matching parameters of the 179 patients with and without DNMT3A/ TET2 CHIP-driver mutations and LOY < 17%. **(D)** Kaplan-Meier survival curves in patients with and without DNMT3A/TET2 CHIP-driver mutations and LOY < 17%.

### Potential mechanistic insights by sc-RNAseq analyses of blood cells

To delineate transcriptional differences in circulating hematopoietic cells with and without LOY and to assess the effect of the presence of CHIP mutations on LOY cells, we additionally analysed scRNA-seq data derived from circulating PBMCs of ten patients with heart failure (mean age of 67.3 years and ejection fraction of 34.4%). ^20^ Among them, four patients did not have CHIP mutations, whereas five patients had DNMT3A CHIP-driver mutations with a VAF ≥ 2%. LOY was defined in scRNA-seq data as combinatorial loss of expression of any gene encoded on the Y chromosome. As illustrated in **Figure 6A**, LOY cells were intermingled with cells expressing genes encoded by the Y chromosome and did not form a specific subcluster. Assessing the relative distribution of LOY cells in different leukocyte subsets revealed that monocytes contribute the highest absolute number of LOY cells, but also are relatively enriched in LOY as compared to other cell types with a mean of 6.99 % LOY cells, whereas LOY cells among T cells showed the lowest absolute and relative contribution (1.64%; **Figure 6B)**. Given the abundance of monocytic LOY cells and the previously reported impact on cardiac fibrosis, we focused on monocytes for further analysis. LOY monocytes showed a significant up-regulation of 193 genes, which were associated with Gene ontology (GO) terms associated with macrophage-mediated tissue damage and cardiac fibrosis such as “Toll Like Receptor 2 (TLR2) Cascade” and “TGF-Beta Signaling Pathway” **(Figure 6C).** Examples of genes enriched in these GO terms are illustrated in the heatmap **(Figure 6D)** demonstrating a pronounced pro-fibrotic, activated gene signature in LOY monocytes. In addition, representative examples of differentially expressed genes in LOY versus Y chromosome carrying cells in the individual patients are shown in **Figure 6E**. Interestingly, we found prototypical profibrotic and tissue-damage associated genes such as the Toll-Like Receptor 2, which promotes cardiac fibrosis via IRAK/NFkB signaling^25^ and *S100A8, CLEC4D* and *USP13,* which augment fibrosis and tissue disruption as members and stabilizers of the damage-associated molecular pattern family. ^47^

**Figure 6.**
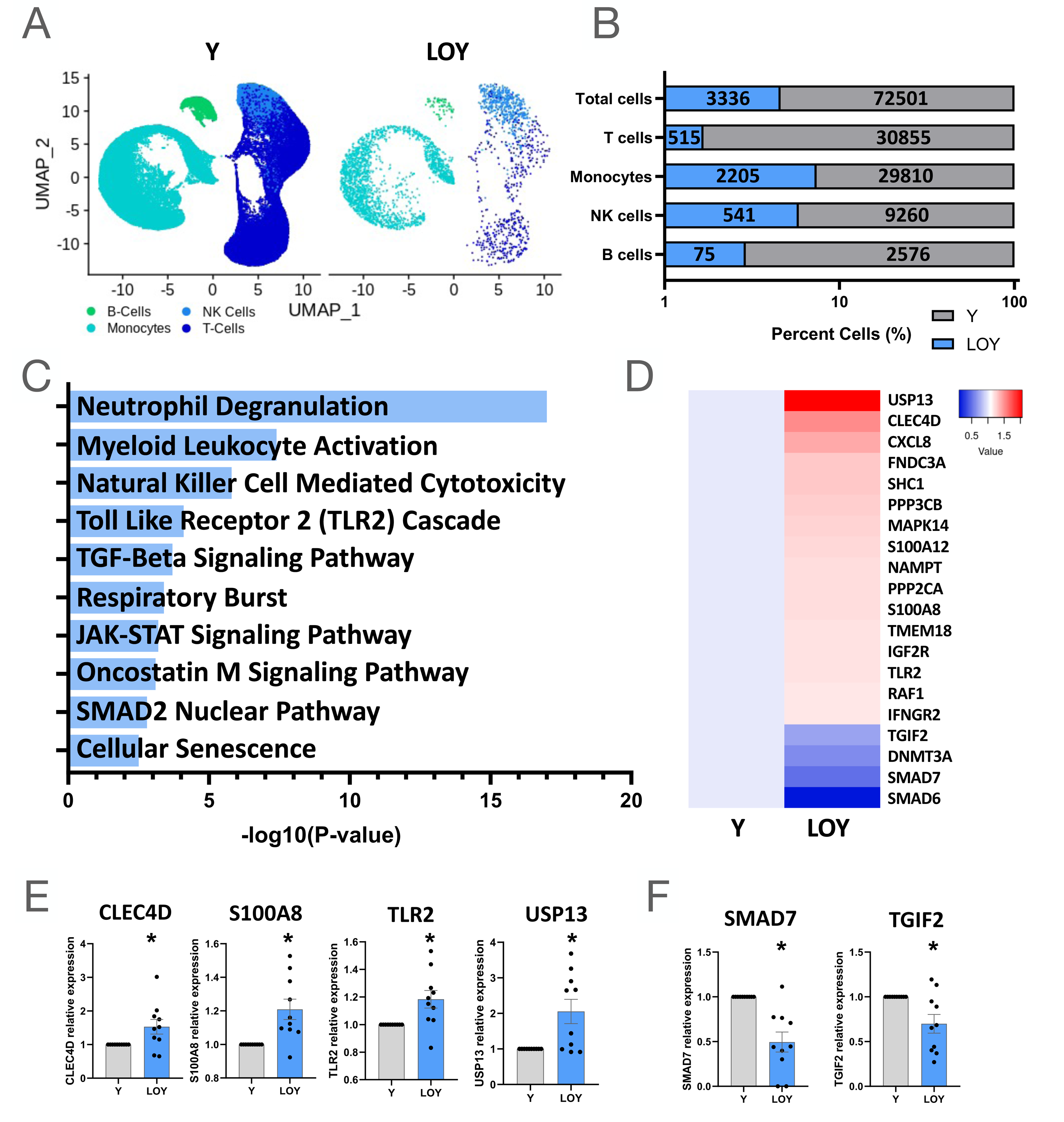
ScRNA-seq analysis of LOY cell-distribution and monocytic profiles in heart failure patients shows pro-fibrotic signature**. (A)** UMAP of PBMCs. **(B)** Percent LOY and Y-harboring cells by cell type with absolute cell counts (n=10). **(C)** GO terms associated with LOY regulated genes. (**D)** Heatmap of regulated genes in LOY monocytes associated with monocyte activation, fibrosis and TGF-b signaling. **(E-F)** Relative expression of **(E)** upregulated and **(F)** downregulated genes by bar plot. Panels A-F: N=10; Significance: p<0.05 by paired *t*-test.

Of the downregulated genes in LOY monocytes, the decreased expression levels of SMAD7 and TGIF2 are of special interest **(Figure 6F),** as both may restrain TGF-ß induced activation pathways. ^45^ Overall, these transcriptional signatures of LOY cells suggest an increased capacity for pro-fibrotic signaling in circulating monocytes lacking Y chromosome encoded genes.

In order to gain further insights into the effect of the combination of both, the presence of harboring CHIP mutations and LOY ≥ 17% at single cell resolution, we conducted a paired analysis of LOY and Y cells in patients with and without DNMT3A mutations. We found unique signatures in LOY cells deriving from patients with or without DNMT3A driver mutations, with only little overlap **(Figure 7A).** Specifically upregulated GO terms in LOY cells derived from patients simultaneously harboring DNMT3A CHIP-driver mutations were related to maladaptive inflammation such as “Regulation of IFNG Signaling”, “Class I MHC Mediated Antigen Processing & Presentation as well as “Toll Like Receptor 2 (TLR2) Cascade” and “Cellular senescence”. In addition, pathways related to fibrosis such as “HIF-1 Signaling Pathway” and “ERK/MAPK Targets” or “Cardiac Hypertrophic Response” were regulated. In contrast, we did not find GO terms related to increased inflammation in LOY cells of patients without CHIP mutations compared to Y cells of the same patients **(Figure 7B).** The differential gene expression associated with LOY in the presence or absence of DNMT3A CHIP driver mutations is further illustrated by the heat map shown in **Figure 7C**. Overall, these data suggests that LOY has a different impact in the presence or absence of DNMT3A CHIP.

**Figure 7:**
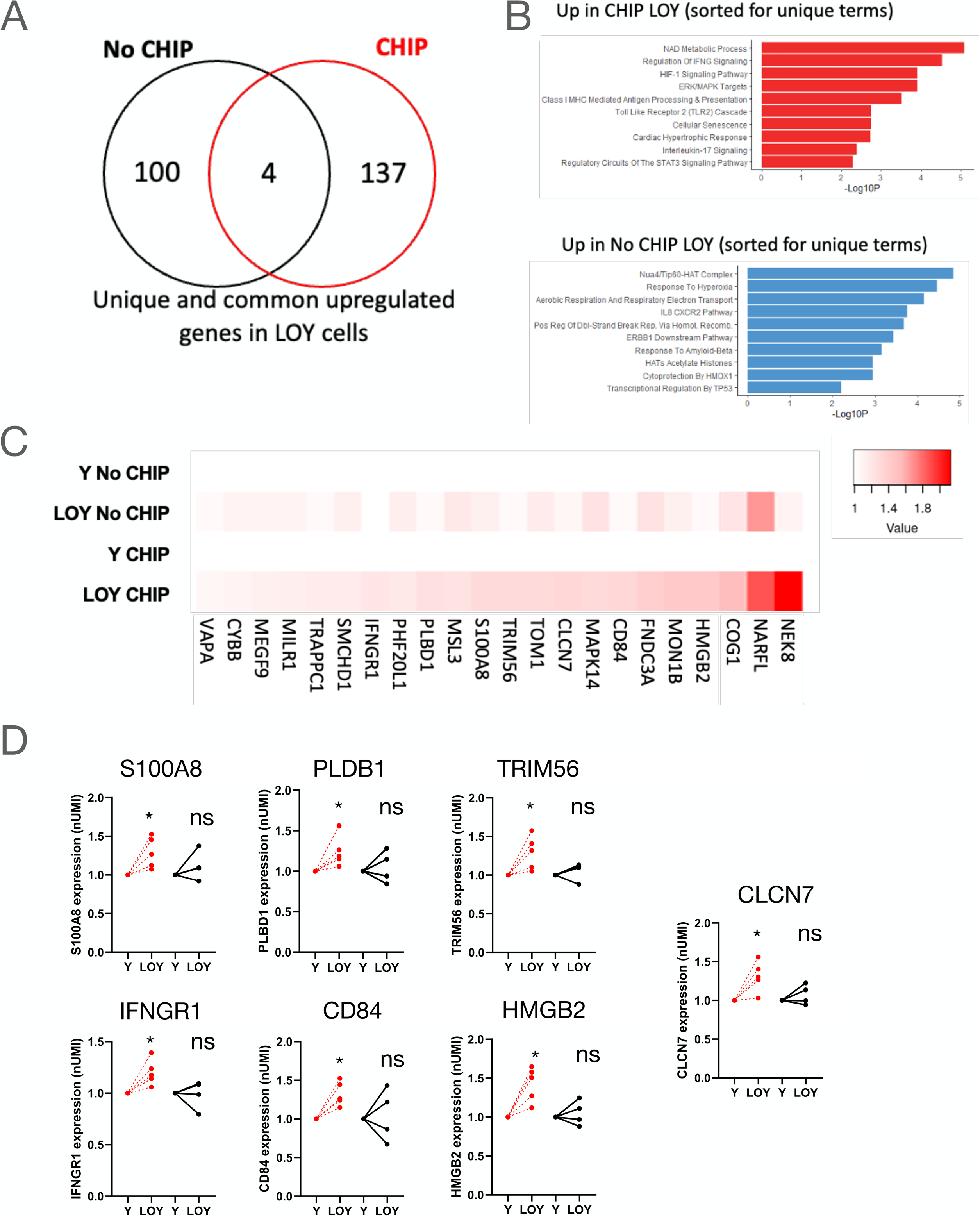
Patient level analysis comparing gene expression in patients with DNMT3A CHIP (n=5) and No CHIP (n=4). (**A)** Venn diagramm displaying common and unique unregulated genes in LOY cells compared to Y cells in patients with CHIP and No CHIP. (**B)** Go Terms associated with LOY regulated genes in patients with CHIP and No CHIP. (**C)** Heatmap of regulated genes in LOY monocytes of patients with CHIP and No CHIP compared to Y cells. (**D)** Patient level analysis of regulated genes in LOY cells compared to Y cells in patients with CHIP and No CHIP. *p<0.05 by paired *t*-test.

This is also reflected at single gene level in LOY cells of patients simultaneously harboring DNMT3A CHIP driver mutations. Here, proinflammatory genes such as the alarmins S100A8, which initiates neutrophil chemotaxis as a damage-associated molecular pattern, and HMGB2, which acts proinflammatory as cytokine in the extracellular space and binds to RAGE to induce oxidative stress and inflammation, were significantly upregulated. Moreover, expression of IFNGR and CD84, which enhances IFN-y secretion, ^26^ as well as the interferon-inducible E3 ubiquitin ligase TRIM56, which modulates cGAS-STING, a critical driver of ageing-related inflammation, were significantly increased.

Finally, PLBD1, which generates lipid mediators of inflammation ^27^, and CLCN7, a major activator of cellular lysosomal activity^28,29^, were significantly augmented only in LOY cells of patients with DNMT3A mutations **(Figure 7D).**

These data support the conclusion that, in addition to profibrotic activation, LOY monocytes of patients simultaneously harboring DNMT3A CHIP-driver mutations display transcriptional changes indicative of augmented generic paracrine and cell-intrinsic inflammation pathways.

## Discussion

The present study is the first to investigate the prognostic significance of LOY, the most common acquired somatic mutation in human blood cells, in patients with established chronic heart failure. Our results provide the following insights: (1) LOY is a major independent determinant of increased mortality. (2) LOY associates with worse outcomes across the entire spectrum of LVEF, thus including both, men with HFrEF as well as with HFpEF. (3) The age-related prevalence of LOY parallels the age-associated prevalence of DNMT3A/TET2 CHIP-driver mutations in chronic heart failure. (4) In men with chronic heart failure, a substantial number of patients harboring CHIP driver mutations also show LOY ≥ 17%. (5) The co-occurrence of LOY in part contributes to the well-established increased mortality observed in patients with chronic heart failure carrying DNMT3A/TET2 CHIP driver mutations. (6) Mechanistically, scRNA-seq analyses of circulating monocytes lacking Y chromosome encoded genes exhibit profibrotic genetic signatures, which are further enhanced by features of augmented generic inflammation pathways in patients simultaneously harboring DNMT3A CHIP-driver mutations. Thus, when addressing the prognostic significance of age-related acquired somatic DNA mutations in circulating blood cells of men with chronic heart failure, it appears to be of utmost importance to account for the presence of LOY.

LOY has been previously linked to shorter life span in elderly men and associated with age-associated degenerative diseases like Alzheimer’s disease, macular degeneration, and cardiovascular disease in large epidemiological studies.^12^ ^30^ ^31^ ^32^ ^33^ It was only until very recently that it was experimentally discovered in a mouse model mimicking LOY in hematopoietic stem cells that LOY led to diffuse cardiac fibrosis during aging, which was accompanied by a progressive decline in cardiac function.^15^ This study indicated for the first time that LOY in hematopoietic cells was linked to heart failure.^34^ Mechanistically, modelling LOY in bone marrow cells led to polarization of bone marrow-derived macrophages lacking the Y chromosome towards a profibrotic phenotype leading to fibroblast activation, deposition of excessive extracellular matrix and cardiac fibrosis in large part mediated via TGF-beta signaling pathways.^15^ We could recently extend these findings into the clinical scenario by studying a very elderly patient population of men undergoing TAVR for severe degenerative aortic valve stenosis.^16^ Using single-cell RNA sequencing analysis of patient derived circulating blood cells, we could demonstrate that LOY in monocytes is associated with a pro-fibrotic gene signature characterized by sensitizing the cells for the TGF-beta signaling pathway.^16^ As diffuse cardiac fibrosis represents the common denominator of progression of all forms of chronic heart failure,^35^ we hypothesized that the presence of LOY might associate with worse survival in patients with established chronic heart failure. Indeed, the results of the present study now disclose that LOY associates with impaired survival of men with chronic heart failure across the entire spectrum of LVEF. Thus, our findings not only further affirm the clinical relevance of the experimental observations by Sano et al.,^15^ but may also open new perspectives on risk factors for worse outcomes as well as precision treatment strategies in men with chronic heart failure. Indeed, the results of the present study further establish a role for profibrotic signaling in LOY monocytes in patients with chronic heart failure suggesting that patients with substantial LOY may derive significant benefit from antifibrotic therapies.

As clonal hematopoiesis, another acquired somatic mosaic condition in circulating myeloid cells, shares many similarities with LOY, including its association with advanced age as well as its prognostic significance in patients with heart failure with reduced ejection fraction,^2–5^ we also investigated a potential interaction between LOY and CHIP in our patient cohort. Mechanistically, DNMT3A/TET2 CHIP driver mutations may exert their detrimental effects on cardiovascular disease predominantly by a proinflammatory activation of circulating blood cells.^36–39^ Specifically, patients with chronic heart failure display a proinflammatory gene signature in their circulating myeloid cells, which is further aggravated in carriers of DNMT3A/TET2 CHIP driver mutations.^40^ In concordance with previous small studies, the prevalence of DNMT3A/TET2 CHIP-driver mutations increased significantly with increasing extent of LOY.^17,41^ Likewise, in carriers of DNMT3A/TET2 CHIP-driver mutations, 24.3 % of patients were simultaneously showing LOY ≥ 17%. Thus, there is considerable coexistence of LOY and CHIP in men with chronic heart failure, which is presumably due to the advanced age of the patients studied, as statistical testing failed to establish a significant association between the presence or extent of LOY and DNMT3A/TET2 driver mutations in our cohort.

Most importantly, the cooccurrence of LOY ≥17% in patients harboring DNMT3A/TET2 CHIP driver mutations in part contributed to the observed prognostic significance of carrying DNMT3A/TET2 CHIP driver mutations, while LOY ≥ 17% was associated with increased mortality during follow up irrespective of the presence of DNMT3A/TET2 CHIP driver mutations. Notably, patients harboring both, LOY ≥ 17% as well as DNMT3A/TET2 CHIP-driver mutations demonstrated the highest HR for mortality. Mechanistically, this observation may be well rationalized by the combination of proinflammatory and profibrotic activation of circulating blood cells contributing to worse clinical outcome in patients with heart failure. ^16 38 37 36 42 43 44^ Indeed, the results of our sc-RNA sequencing analysis extended our previously published observation in elderly patients undergoing TAVR^16^ by demonstrating that LOY in circulating monocytes of patients with chronic heart failure associates with an increased profibrotic gene signature. Specifically, expression levels of TGIF2 and SMAD7, which restrain TGF-ß induced activation pathways and prevent TGFß-mediated fibrosis in postinfarction failing hearts,^45^ were significantly downregulated, while expression of TLR2 and S100A8, which promote inflammation, fibrosis and progression of heart failure, ^25,46,47^ were significantly induced in LOY monocytes.

More importantly, LOY monocytes in patients simultaneously harboring DNMT3A CHIP-driver mutations demonstrated a boosted, more generic inflammatory activation genetic signature compared to LOY monocytes of patient harboring no CHIP-driver mutations. Here, expression of HMGB2 and PLBD1, which act proinflammatory in a paracrine fashion either by acting as cytokine itself of by generating lipid mediators of inflammation, respectively, and were both shown to correlate with cardiac functional deterioration post-MI^48,27,49^ were specifically upregulated in LOY monocytes of patients simultaneously harboring DNMT3A CHIP-driver mutations. Moreover, expression of TRIM56 and IFNGR1, which both act as mediators of immune dysregulation by activating the cGAS-STING signaling pathway,^50,51^ a critical driver of ageing-related inflammation^52^ and known to contribute to adverse left ventricular remodeling leading to heart failure in mice,^53^ were augmented in LOY monocytes of patients with DNMT3A CHIP-driver mutations.

In conclusion, the complex relationship between different classes of age-related somatically acquired mutations needs to be considered when assessing clinical outcomes. At least for patients with chronic heart failure, LOY ≥17% appears to be an independent risk factor for increased mortality, and its co-occurrence with CHIP may partly confound the prognostic significance of DNMT3A/TET2 CHIP driver mutations.

### Limitations

The present clinical study cannot provide a cause-and-effect relationship between LOY and increased mortality in patients with chronic heart failure. As such, we can only conclude that LOY is an important independent predictor of mortality. In addition, we can only report all-cause mortality data, but cannot provide data on cause specific mortality or hospitalizations for heart failure. Moreover, our findings may only relate to patients with established chronic heart failure but may not be applicable to other forms of cardiovascular disease or incident heart failure. In addition, the patients with heart failure and preserved ejection fraction of the present study do not represent the classical HFpEF population, but rather refer in large part to patients with persistent symptoms and signs of heart failure secondary to aortic stenosis induced left ventricular hypertrophy even after successful removal of the stenotic valve by TAVR. However, subanalysis excluding patients post TAVR confirmed the prognostic significance of LOY. Nevertheless, if further studies in even larger cohorts would confirm the present results, LOY may not only provide for risk stratification, but also for precision tailored therapies in men with chronic heart failure across the entire spectrum of ejection fraction. Second, we used an Youden-Index derived optimal cut-off of 17% to assess the effects of LOY on mortality. As in our previous report in elderly patients with severe aortic valve stenosis undergoing TAVR,^16^ there was a dose-dependent increase in mortality with increasing extent of LOY with a sharp increase in the hazard ratios for death at LOY ≥17%. In contrast, for DNMT3A/TET2 mutations, we did not select an optimized cut-off level, but rather used the generally accepted definition of CHIP as ≥2% VAF. Moreover, our scRNAseq analyses of circulating monocytes only refer to LOY in patients with or without simultaneously harboring DNMT3A CHIP-driver mutations. Thus, although it has been recently reported that the enhanced inflammation in loss of Dnmt3a in myeloid cells experimentally phenocopies the effect of loss of Tet2 in a mouse model of atherosclerosis^43^, we cannot comment on potential combinatorial effects of LOY and TET2 CHIP-driver mutations in patients with chronic heart failure. Third, we primarily focused on the two most common CHIP-driver mutations, DNMT3A and TET2, for which a prognostic role in heart failure is firmly established. Thus, we cannot comment on other pathogenic CHIP driver mutations implicated in cardiovascular diseases. Finally, mortality rates were significantly lower in our validation cohort, which may have affected the statistical significance of the analysis. However, hazard ratios for the prognostic relevance of harboring LOY were comparable in the initial cohort and in the validation cohort.

## Data Availability

All data produced in the present study are available upon reasonable request to the authors

---

**Figure S1.**
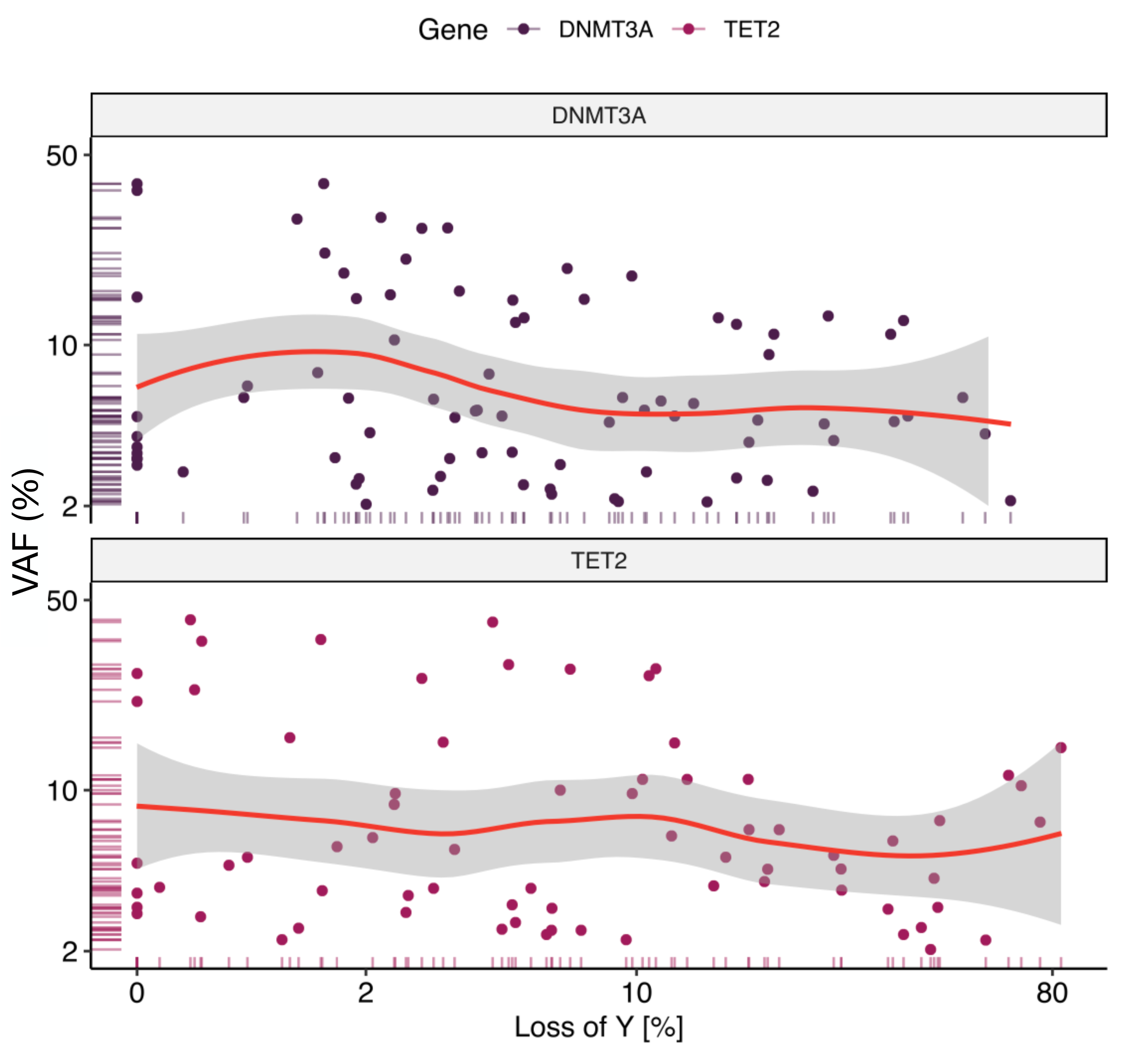
Correlation plots of LOY and VAF of DNMT3A CHIP-driver mutations (upper panel) and TET2 CHIP-driver mutations (lower panel). The red line shows locally estimated scatterplot smoothing with 95% confidence intervals.

**Figure S2.**
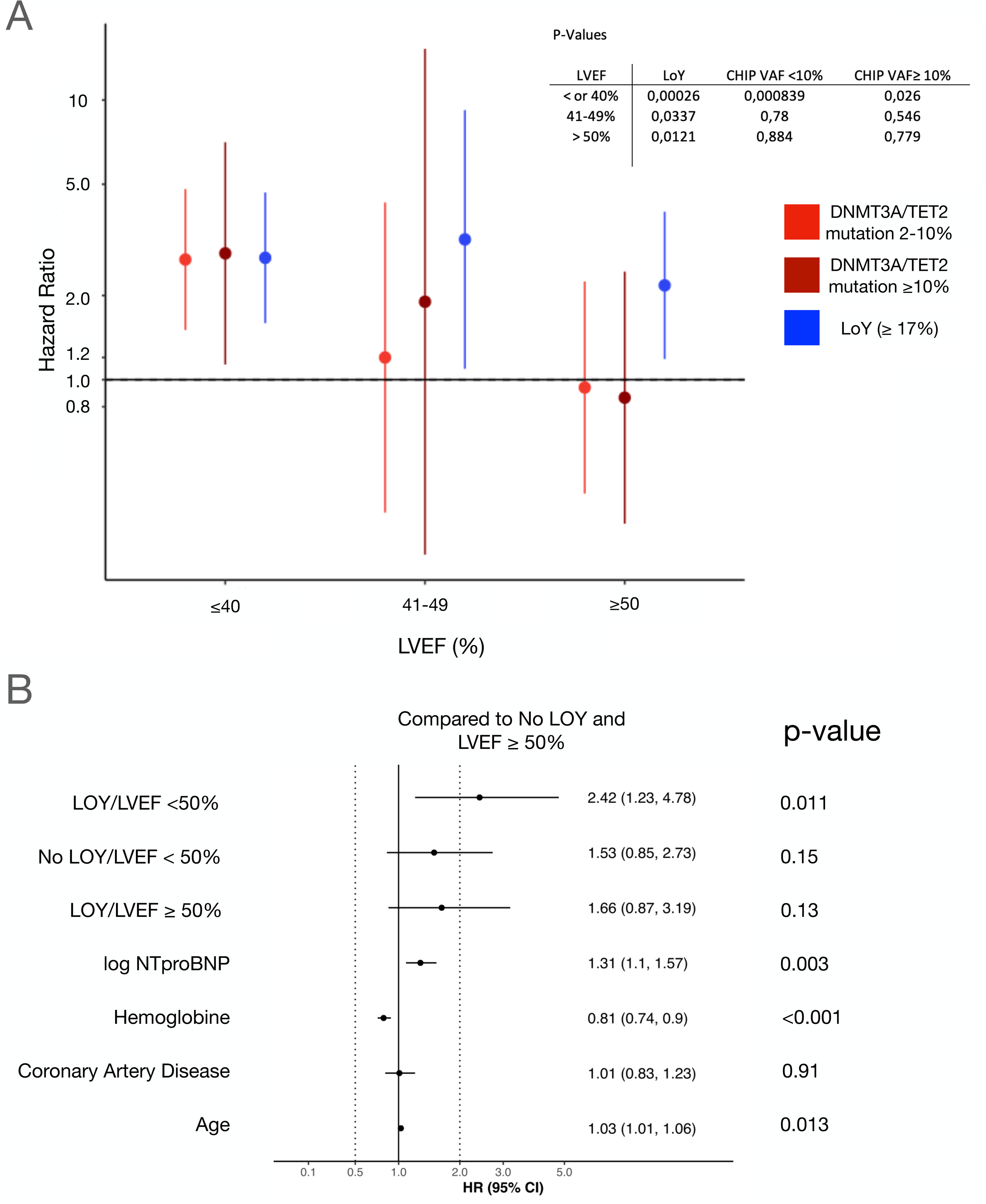
**(A)** Effect of harboring LOY or DNMT3A/TET2 CHIP-driver mutations on all-cause mortality in 3 groups: men with reduced (≤40%), mildly reduced (41-49%) and preserved (≥50%) ejection fraction. Dots represent hazard ratios (HR), lines represent 95% confidence intervals (CI). DNMT3A/TET2 CHIP driver mutations are separated in to large (VAF >10%) and small clones (VAF 2-10%). **(B)** Multivariate analysis reveals that the risk of death in patients with heart failure and LOY ≥17% is highest in patients with reduced ejection fraction.

**Table S1:** Variants of DNMT3A mutations in the present study.

| Study ID | HUGO Symbol | Chromosome | VAF (%) | DNA Change | Protein Change | Variant |
| --- | --- | --- | --- | --- | --- | --- |
| 224 | TET2 | 4 | 2,04 | c.944_947del | p.Ser315Phefs*31 | DEL |
| 20 | TET2 | 4 | 2,28 | c.2771del | p.His924Leufs*29 | DEL |
| 290 | TET2 | 4 | 2,29 | c.4708del | p.Met1570* | DEL |
| 506 | TET2 | 4 | 2,29 | c.4082G>A | p.Gly1361Asp | PM |
| 12 | TET2 | 4 | 2,43 | c.4210C>T | p.Arg1404* | PM |
| 299 | TET2 | 4 | 2,43 | c.2743dup | p.Ala915Glyfs*9 | DUP |
| 222 | TET2 | 4 | 2,55 | c.2279_2280del | p.Phe760Serfs*8 | DEL |
| 43 | TET2 | 4 | 2,55 | c.5482C>T | p.Gln1828* | PM |
| 270 | TET2 | 4 | 2,58 | c.3893G>A | p.Cys1298Tyr | PM |
| 353 | TET2 | 4 | 2,61 | c.724G>C | p.Val242Leu | PM |
| 219 | TET2 | 4 | 2,63 | c.3895A>G | p.Lys1299Glu | PM |
| 273 | TET2 | 4 | 2,78 | c.5618T>C | p.Ile1873Thr | PM |
| 237 | TET2 | 4 | 2,96 | c.5618T>C | p.Ile1873Thr | PM |
| 298 | TET2 | 4 | 3,06 | c.4060dup | p.Arg1354Lysfs*47 | DUP |
| 379 | TET2 | 4 | 3,10 | c.76C>T | p.Gln26* | PM |
| 145 | TET2 | 4 | 3,21 | c.822del | p.Asn275Ilefs*18 | DEL |
| 318 | TET2 | 4 | 3,24 | c.3848C>A | p.Ala1283Asp | PM |
| 5 | TET2 | 4 | 3,27 | c.2652del | p.Glu885Asnfs*36 | DEL |
| 549 | TET2 | 4 | 3,27 | c.4133G>A | p.Cys1378Tyr | PM |
| 307 | TET2 | 4 | 3,36 | c.2266dup | p.Ile756Asnfs*13 | DUP |
| 378 | TET2 | 4 | 3,70 | c.4626_4627insTAG | p.Gln1542* | INS |
| 185 | TET2 | 4 | 3,79 | c.5618T>C | p.Ile1873Thr | PM |
| 557 | TET2 | 4 | 3,89 | c.5189_5192dup | p.His1732Trpfs*24 | DUP |
| 562 | TET2 | 4 | 3,91 | c.3646C>T | p.Arg1216* | PM |
| 119 | TET2 | 4 | 3,98 | c.500del | p.Ser167* | DEL |
| 175 | TET2 | 4 | 3,98 | c.4894C>T | p.Gln1632* | PM |
| 133 | TET2 | 4 | 4,02 | c.4460del | p.Asn1487Ilefs*84 | DEL |
| 135 | TET2 | 4 | 4,08 | c.4031C>T | p.Ala1344Val | PM |
| 176 | TET2 | 4 | 4,26 | c.4072T>C | p.Cys1358Arg | PM |
| 180 | TET2 | 4 | 4,40 | c.860_867del | p.Lys287Serfs*5 | DEL |
| 82 | TET2 | 4 | 4,81 | c.3581C>G | p.Pro1194Arg | PM |
| 376 | TET2 | 4 | 4,82 | c.1651del | p.Asp551Ilefs*10 | DEL |
| 380 | TET2 | 4 | 5,00 | c.1842del | p.Leu615Serfs*24 | DEL |
| 162 | TET2 | 4 | 5,10 | c.3932T>C | p.Leu1311Pro | PM |
| 271 | TET2 | 4 | 5,40 | c.1607_1608del | p.Lys536Argfs*30 | DEL |
| 363 | TET2 | 4 | 5,40 | c.1777C>T | p.Gln593* | PM |
| 156 | TET2 | 4 | 5,50 | c.2961C>A | p.Cys987* | PM |
| 580 | TET2 | 4 | 5,82 | c.2150_2151del | p.His717Profs*5 | DEL |
| 583 | TET2 | 4 | 5,97 | c.3643G>T | p.Glu1215* | PM |
| 230 | TET2 | 4 | 6,30 | c.3872G>A | p.Trp1291* | PM |
| 295 | TET2 | 4 | 6,50 | c.5718_5722dup | p.Asn1908Thrfs*2 | DUP |
| 308 | TET2 | 4 | 6,60 | c.3487del | p.Ile1163Serfs*63 | DEL |
| 592 | TET2 | 4 | 6,99 | c.3814G>C | p.Ala1272Pro | PM |
| 606 | TET2 | 4 | 7,00 | c.3734A>G | p.Tyr1245Cys | PM |
| 14 | TET2 | 4 | 7,50 | c.553C>T | p.Gln185* | PM |
| 141 | TET2 | 4 | 7,60 | c.4256C>G | p.Pro1419Arg | PM |
| 256 | TET2 | 4 | 8,80 | c.3500+2T>G | p.splice site mutation | PM |
| 76 | TET2 | 4 | 9,70 | c.1930C>T | p.Gln644* | PM |
| 352 | TET2 | 4 | 9,70 | c.4210C>T | p.Arg1404* | PM |
| 15 | TET2 | 4 | 10,00 | c.1971dup | p.His658Thrfs*23 | DUP |
| 344 | TET2 | 4 | 10,40 | c.5581G>A | p.Gly1861Arg | PM |
| 64 | TET2 | 4 | 11,00 | c.4133G>A | p.Cys1378Tyr | PM |
| 138 | TET2 | 4 | 11,00 | c.3803+2T>C | p.splice site mutation | PM |
| 252 | TET2 | 4 | 11,00 | c.369dup | p.Asn124* | DUP |
| 306 | TET2 | 4 | 11,40 | c.3655C>G | p.His1219Asp | PM |
| 127 | TET2 | 4 | 14,50 | c.3578G>A | p.Cys1193Tyr | PM |
| 369 | TET2 | 4 | 15,10 | c.2474C>G | p.Ser825* | PM |
| 286 | TET2 | 4 | 15,20 | c.3190del | p.Val1064Phefs*2 | DEL |
| 169 | TET2 | 4 | 15,80 | c.3384_3385insTT | p.Asp1129Leufs*9 | INS |
| 254 | TET2 | 4 | 21,50 | c.3501-1G>A | p.splice site mutation | PM |
| 651 | TET2 | 4 | 23,73 | c.3594+1G>C | p.splice site mutation | PM |
| 250 | TET2 | 4 | 26,10 | c.2150_2151del | p.His717Profs*5 | DEL |
| 71 | TET2 | 4 | 26,70 | c.4120T>C | p.Cys1374Arg | PM |
| 350 | TET2 | 4 | 27,20 | c.1747G>T | p.Glu583* | PM |
| 80 | TET2 | 4 | 28,20 | c.4707C>G | p.Tyr1569* | PM |
| 136 | TET2 | 4 | 28,30 | c.4073G>A | p.Cys1358Tyr | PM |
| 236 | TET2 | 4 | 29,30 | c.4108G>A | p.Gly1370Arg | PM |
| 383 | TET2 | 4 | 35,60 | c.818del | p.Gln273Argfs*20 | DEL |
| 347 | TET2 | 4 | 36,10 | c.2839C>T | p.Gln947* | PM |
| 181 | TET2 | 4 | 41,70 | c.3721G>A | p.Ala1241Thr | PM |
| 231 | TET2 | 4 | 42,50 | c.5158A>T | p.Lys1720* | PM |

PM - point mutation, DEL - deletion, DUP - duplication, INS - insertion, COMP - complex mutation

**Table S2:** Variants of TET2 mutations in the present study.

PM - point mutation, DEL - deletion, DUP - duplication, INS - insertion, COMP - complex mutation
| StudyID | HUGO Symbol | Chromosome | VAF (%) | DNA Change | Protein Change | Variant |
| --- | --- | --- | --- | --- | --- | --- |
| 600 | DNMT3A | 2 | 2,04 | c.2245C>G | p.Arg749Gly | PM |
| 98 | DNMT3A | 2 | 2,10 | c.1954del | p.Asp652Thrfs*53 | DEL |
| 605 | DNMT3A | 2 | 2,11 | c.677C>T | p.Ala226Val | PM |
| 147 | DNMT3A | 2 | 2,13 | c.1585G>A | p.Asp529Asn | PM |
| 615 | DNMT3A | 2 | 2,18 | c.2026C>T | p.Arg676Trp | PM |
| 317 | DNMT3A | 2 | 2,30 | c.1006T>A | p.Phe336Ile | PM |
| 282 | DNMT3A | 2 | 2,38 | c.1662C>A | p.Cys554* | PM |
| 633 | DNMT3A | 2 | 2,41 | c.1345_1348del | p.Ala449delfs | DEL |
| 323 | DNMT3A | 2 | 2,44 | c.1851+1G>T | p.splice site mutation | PM |
| 226 | DNMT3A | 2 | 2,56 | c.2132_2135dup | p.Asp712Glufs*2 | DUP |
| 69 | DNMT3A | 2 | 2,58 | c.1856del | p.Pro619Leufs*32 | DEL |
| 124 | DNMT3A | 2 | 2,69 | c.1843C>T | p.Gln615* | PM |
| 642 | DNMT3A | 2 | 2,74 | c.748C>T | p.Pro250Ser | PM |
| 215 | DNMT3A | 2 | 2,76 | c.2644C>T | p.Arg882Cys | PM |
| 257 | DNMT3A | 2 | 2,81 | c.954_961del | p.Arg320* | DEL |
| 223 | DNMT3A | 2 | 2,95 | c.1609_1613del | p.Cys537Hisfs*7 | DEL |
| 288 | DNMT3A | 2 | 2,95 | c.896A>T | p.Lys299Ile | PM |
| 645 | DNMT3A | 2 | 3,17 | c.2165del | p.Gly722Alafs | DEL |
| 15 | DNMT3A | 2 | 3,19 | c.1851+1delinsAA | p.splice site mutation | COMP |
| 266 | DNMT3A | 2 | 3,39 | c.2711C>T | p.Pro904Leu | PM |
| 212 | DNMT3A | 2 | 3,40 | c.905G>T | p.Gly302Val | PM |
| 397 | DNMT3A | 2 | 3,40 | c.2644C>T | p.Arg882Cys | PM |
| 8 | DNMT3A | 2 | 3,43 | c.2193_2195del | p.Phe732del | DEL |
| 650 | DNMT3A | 2 | 3,59 | c.1713_1718del | p.Ala572_Gln573del | DEL |
| 104 | DNMT3A | 2 | 3,61 | c.2207G>A | p.Arg736His | PM |
| 307 | DNMT3A | 2 | 3,63 | c.1397_1399delinsC | p.Lys466Thrfs*6 | COMP |
| 162 | DNMT3A | 2 | 3,83 | c.1667+1G>T | p.splice site mutation | PM |
| 652 | DNMT3A | 2 | 4,02 | c.1097G>A | p.Arg366His | PM |
| 156 | DNMT3A | 2 | 4,09 | c.2207G>A | p.Arg736His | PM |
| 653 | DNMT3A | 2 | 4,25 | c.1904G>A | p.Arg635Gln | PM |
| 177 | DNMT3A | 2 | 4,37 | c.915G>A | p.Trp305* | PM |
| 313 | DNMT3A | 2 | 4,42 | c.2221dup | p.Ala741Glyfs*8 | DUP |
| 97 | DNMT3A | 2 | 4,82 | c.2578T>C | p.Trp860Arg | PM |
| 274 | DNMT3A | 2 | 4,90 | c.2578T>C | p.Trp860Arg | PM |
| 39 | DNMT3A | 2 | 4,93 | c.2173+2T>A | p.splice site mutation | PM |
| 247 | DNMT3A | 2 | 5,00 | c.2210T>G | p.Leu737Arg | PM |
| 658 | DNMT3A | 2 | 5,13 | c.2638A>G | p.Met880Val | PM |
| 662 | DNMT3A | 2 | 5,17 | c.841G>T | p.Glu281* | PM |
| 30 | DNMT3A | 2 | 5,20 | c.2478+1G>A | p.splice site mutation | PM |
| 270 | DNMT3A | 2 | 5,20 | c.2363C>T | p.Ala788Val | PM |
| 366 | DNMT3A | 2 | 5,20 | c.1969G>A | p.Val657Met | PM |
| 459 | DNMT3A | 2 | 5,46 | c.2082+1G>A | p.splice site mutation | PM |
| 204 | DNMT3A | 2 | 5,50 | c.2032C>T | p.Gln678* | PM |
| 349 | DNMT3A | 2 | 5,50 | c.2311C>T | p.Arg771* | PM |
| 665 | DNMT3A | 2 | 5,86 | c.2206C>T | p.Arg736Cys | PM |
| 355 | DNMT3A | 2 | 6,00 | c.976C>T | p.Arg326Cys | PM |
| 6 | DNMT3A | 2 | 6,10 | c.2083-2A>G | p.splice site mutation | PM |
| 666 | DNMT3A | 2 | 6,16 | c.2536C>T | p.Gln846* | PM |
| 173 | DNMT3A | 2 | 6,20 | c.792del | p.Val265Trpfs*51 | DEL |
| 377 | DNMT3A | 2 | 6,20 | c.1752C>A | p.Tyr584* | PM |
| 669 | DNMT3A | 2 | 6,20 | c.2330C>A | p.Pro777His | PM |
| 271 | DNMT3A | 2 | 6,90 | c.1572T>A | p.Cys524* | PM |
| 248 | DNMT3A | 2 | 7,70 | c.2153del | p.Pro718Leufs*61 | DEL |
| 22 | DNMT3A | 2 | 7,80 | c.1976G>A | p.Arg659His | PM |
| 673 | DNMT3A | 2 | 9,18 | c.427C>T | p.Arg143* | PM |
| 681 | DNMT3A | 2 | 10,46 | c.2390A>G | p.Asn797Ser | PM |
| 108 | DNMT3A | 2 | 11,00 | c.2645G>A | p.Arg882His | PM |
| 115 | DNMT3A | 2 | 11,00 | c.1481G>T | p.Cys494Phe | PM |
| 149 | DNMT3A | 2 | 12,00 | c.1961G>T | p.Gly654Val | PM |
| 273 | DNMT3A | 2 | 12,20 | c.2446C>T | p.Gln816* | PM |
| 12 | DNMT3A | 2 | 12,40 | c.2679G>A | p.Trp893* | PM |
| 228 | DNMT3A | 2 | 12,70 | c.2645G>A | p.Arg882His | PM |
| 289 | DNMT3A | 2 | 12,70 | c.2644C>T | p.Arg882Cys | PM |
| 220 | DNMT3A | 2 | 12,90 | c.1733A>G | p.Glu578Gly | PM |
| 48 | DNMT3A | 2 | 14,80 | c.1549T>C | p.Cys517Arg | PM |
| 685 | DNMT3A | 2 | 14,91 | c.995G>A | p.Gly332Glu | PM |
| 294 | DNMT3A | 2 | 15,00 | c.1902_1904dup | p.Arg635dup | DUP |
| 331 | DNMT3A | 2 | 15,20 | c.2716A>G | p.Lys906Glu | PM |
| 109 | DNMT3A | 2 | 15,50 | c.2645G>A | p.Arg882His | PM |
| 66 | DNMT3A | 2 | 16,00 | c.2174-2A>G | p.splice site mutation | PM |
| 150 | DNMT3A | 2 | 18,20 | c.1610G>T | p.Cys537Phe | PM |
| 686 | DNMT3A | 2 | 18,65 | c.1106T>G | p.Ile369Ser | PM |
| 102 | DNMT3A | 2 | 19,40 | c.2409-2A>C | p.splice site mutation | PM |
| 379 | DNMT3A | 2 | 21,00 | c.1135C>G | p.Arg379Gly | PM |
| 359 | DNMT3A | 2 | 22,10 | c.2644C>T | p.Arg882Cys | PM |
| 324 | DNMT3A | 2 | 27,20 | c.2645G>A | p.Arg882His | PM |
| 284 | DNMT3A | 2 | 27,30 | c.1474+1G>A | p.splice site mutation | PM |
| 267 | DNMT3A | 2 | 29,40 | c.1906G>A | p.Val636Met | PM |
| 186 | DNMT3A | 2 | 29,80 | c.2644C>T | p.Arg882Cys | PM |
| 129 | DNMT3A | 2 | 37,30 | c.2159G>A | p.Arg720His | PM |
| 50 | DNMT3A | 2 | 39,40 | c.2711C>T | p.Pro904Leu | PM |
| 691 | DNMT3A | 2 | 39,45 | c.2099C>T | p.Pro700Leu | PM |

PM - point mutation, DEL - deletion, DUP - duplication, COMP - complex mutation

## Acknowledgements

S.C. is supported by the DFG (SFB1531, project code 456687919 and FOR 5643, project code 515629962). M.S. is supported by an excellence grant of the German Center for Cardiovascular Research (DZHK-81X3600506), the German CHIP Registry (Deutsches CHIP Register e.V.), and a Junior Research Group Cardiovascular Diseases Grant of the CORONA Foundation (S199/10085/2021). S.M.-P. received support by a Cardiopulmonary Institute (CPI) Excellence Cluster Start-up Grant and a Center for Cardiovascular Research (DZHK) Postdoc Start-up Grant and the DFG FOR 5643, project code 515629962). T.S. is supported by the Else Kroener-Fresenius Foundation and DFG (SFB TRR 219, Project-ID 322900939). S.D. is supported by the DFG (SFB1531, project code 456687919 and FOR 5643, project code 515629962). AMZ is funded/co-funded by the European Union (ERC-2021-ADG, GAP - 101054899, CHIP AVS) and the DFG (FOR 5643, project code 515629962). WA is funded by the DFG (FOR 5643, project code 515629962). We would like to thank Dr. Gregor Hoermann, Dr. Manja Meggendorfer and Prof. Torsten Haferlach for their support from the Munich Leukemia Laboratory (MLL).

Views and opinions expressed are however those of the author(s) only and do not necessarily reflect those of the European Union or the European Research Council. Neither the European Union nor the granting authority can be held responsible for them.

## Contributions

K.K. and J.K. performed dataset creation and annotation. S.M.P collected data. T.R. performed LOY measurements. L.T., M.S., W.A and T.S. performed statistical analysis and interpreted data. D.L., H.S. and S.D. advised on the scientific direction of the study and interpreted data. S.C., M.S. and A.M.Z. supervised dataset creation and annotation, interpreted data and oversaw statistical analysis. S.C. and A.M.Z. conceptualized analysis plan. A.M.Z. conceived the study, secured funding and supervised the research. S.C. and A.M.Z. drafted the manuscript. All authors critically revised the manuscript for important intellectual content. All authors provided their final approval of the version to be published.

## Notes

### Competing Interest Statement

The authors have declared no competing interest.

### Author Declarations

Ethics committee/IRB of the Goethe University, Frankfurt Medical School gave ethical approval for this work

